# Examining Australian public perceptions and behaviors towards a future COVID-19 vaccine

**DOI:** 10.1101/2020.09.29.20204396

**Authors:** Holly Seale, Anita E Heywood, Julie Leask, Meru Sheel, David N Durrheim, Katarzyna Bolsewicz, Rajneesh Kaur

**Affiliations:** School of Population Health, University of New South Wales, Sydney, NSW, Australia; Susan Wakil School of Nursing and Midwifery, University of Sydney, Sydney, NSW, Australia; National Centre for Immunisation Research and Surveillance, Kids Research, Sydney Children’s Hospitals Network, Westmead, NSW, Australia; National Centre for Epidemiology and Population Health, Research School of Population Health, ANU College of Health and Medicine, The Australian National University, Acton, ACT, Australia; School of Medicine and Public Health, University of Newcastle, Wallsend, NSW, Australia; Office of Medical Education, Faculty of Medicine, University of New South Wales, Sydney, NSW, Australia

**Author notes:** Correspondence Dr. Holly Seale, School of Population Health, Level 2, Samuels Building, Faculty of Medicine, UNSW Australia, Sydney 2052, Australia.

**Keywords:** immunisation, vaccination decisions, COVID-19, pandemic, acceptance, attitudes, communication

## Abstract

**Background:** There is an indication that vaccine(s) for COVID-19 could be available by early 2021. As immunisation program launches have previously demonstrated, it is essential that careful planning occurs now to ensure the readiness of the public for a COVID-19 vaccine. As part of that process, this study aimed to understand the public perceptions regarding a future COVID-19 vaccine in Australia.

**Methods:** A national cross-sectional online survey of 1420 Australian adults (18 years and older) was undertaken between 18 and 24 March 2020. The statistical analysis of the data included univariate and multivariate logistic regression analysis.

**Results:** Participants generally held positive views towards vaccination. Eighty percent (n=1143) agreed with the statement that *getting myself vaccinated for COVID-19 would be a good way to protect myself against infection*. Females (614, 83%) were more likely to agree with the statement than males (529, 78%) (aOR=1.4 (95% CI: 1.1-1.8); P=0.029), while 90.9% aged 70 and above agreed compared to 76.6% aged 18-29 year old (aOR=2.3 (95% CI:1.2-4.1); 0.008). Agreement was also higher for those with a self-reported chronic disease (aOR=1.4 (95% CI: 1.1-2.0); P=0.043) and among those who held private health insurance (aOR=1.7 (95% CI: 1.3-2.3); P<0.001). Beyond individual perceptions, 78% stated that their decision to vaccinate would be supported by family and friends

**Conclusion:** This study presents an early indication of public perceptions towards a future COVID-19 vaccine and represents a starting point for mapping vaccine perceptions. To support an effective launch of these new vaccines, governments need to use this time to understand the communities concerns and to identify the strategies that will support engagement.

## BACKGROUND

Finding safe and effective vaccine candidates to control the spread of SARS-CoV-2 (COVID-19) is an urgent public health priority. There are an unprecedented number of agencies (including biotechnology companies, universities, military researchers, and pharmaceutical companies) aiming to identify and develop a vaccine candidate at a speed and scale not previously seen [1, 2]. While a smaller number of entities have already launched clinical trials, it is suggested that a COVID-19 vaccine will take 12 to 18 months to develop and manufacture at scale, and may be ready by early 2021 [3].

To ensure community readiness, it is essential that governments determine levels of demand and acceptance of the COVID-19 vaccine to ensure the readiness of both the public and healthcare providers for a COVID-19 vaccine. It is likely that controlling COVID-19 with vaccination will require a critical proportion of the population to accept and receive the vaccine. A minimal target level may exceed 70% accounting for vaccine effectiveness and mechanism of protection, the size of the population in which the vaccine is contraindicated and other factors. However, having a COVID-19 vaccine available does not necessarily equate to people accepting it, as history demonstrates. For example, compliance with the influenza pandemic specific vaccine in 2009 was low, despite higher levels of reported ‘willingness to vaccinate’, which highlights the challenges with compliance and acceptance [4, 5]. To support the launch of a COVID-19 vaccine program and to ensure that communication efforts are attuned to factors impacting on acceptance, we sought to understand people’s attitudes towards vaccination against COVID-19.

## METHODS

The methods used for this study have been previously published [6]. In summary, an online survey of Australian residents was undertaken via a market research company (Quality Online Research (QOR)) between 18 and 24 March 2020. A sample size of 1400 provided us with a sample error of ±3%. Proportional quota sampling was used to ensure that respondents were demographically representative of the Australian public, with quotas based on age, gender, and state/territory. Respondents were required to be 18 years or older and to speak English. After reading the participant information, consent was implied if the person completed the survey and submitted it via the QOR website. Ethics approval for the study was obtained from the University of New South Wales (HC200190).

The questions for this survey were adapted from published studies by HS during the 2009 influenza H1N1/A pandemic [4, 7]. The study tool is available upon request. Questions captured: (1) perceptions of the effectiveness of vaccines in general; (2) priorities for COVID-19 vaccine roll out; and (3) social influences. As a measure of vaccine acceptance, participants were asked if they agreed or disagreed with the following statement: ‘*Getting myself vaccinated for COVID-19 would be a good way to protect myself against infection’*. This item was measured on a 5-point Likert scale with 1=strongly disagree to 5=strongly agree. This variable was treated as the primary outcome with responses collapsed into strongly disagree/disagree/neutral=0 and agree/strongly agree=1. Risk perception of COVID-19 infection was measured on a scale of 1-5 with 1=low risk and 5=very high risk. Lastly data was collected on gender, age, education and employment status, children (including attendance at childcare/school), country of birth/language spoken at home, whether they identified as Aboriginal and/or Torres Strait Islander, international travel patterns since 1 January 2020, private healthcare insurance coverage, income protection insurance, the presence of any chronic illness and self-reported health status (very good, good, moderate, poor, very poor). Due to the uncertainty around vaccine development at the time of the survey, participants were not directly asked whether they would receive a vaccine but rather whether they thought a COVID-19 vaccine would be a good way to protect against infection.

Descriptive statistical statistics were reported for sample demographics. Mean scores and standard deviations of the risk perception score and the vaccine acceptance response were calculated by demographic characteristic. Univariate associations were ascertained with each demographic variable and the outcome variable, vaccine acceptance. The risk perception score of those who would accept the vaccine was compared to those who would not using an independent samples t-test and ANOVA with Bonferroni correction. A multivariate logistic regression model was created with backward elimination model selection and a threshold P value of 0.25 for inclusion of predictor variables. For all analyses, P values of less than 0.05 were considered statistically significant. Data were analyzed using the SPSS software version 26.0 (SPSS Science, Chicago, IL, USA).

## RESULTS

The demographic characteristics of the 1420 respondents by their risk perception and stated vaccine acceptance are presented in Table 1. In summary, 681 (48%) were male, 829 (58%) were in some form of employment, 363 (25%) had a chronic health condition, while 830 (58%) had private health insurance. Participants generally held positive views towards vaccination, with 1188 (83%) agreeing with the statement that ‘vaccines are effective at preventing diseases’, while 305 (21%) indicated that ‘diseases provide better immunity than vaccines do’. Among all respondents, 88% (n=1252) had heard that a COVID-19 vaccine was being developed. Of those who were not aware, 129/168 (76%) were aged under 50 years (lowest awareness levels were in the youngest age group i.e. 18-29 years (n=62/168, 36.9%)). 1195/1420 (84%) agreed that they generally do what their healthcare professional recommends.

**Table 1.** **Covid-19 risk perception across sociodemographic characteristics**

|  | **Total**<br>**n=1420**<br>**n (%)** | **Risk**<br>**perception**<br>**score**<br>**Mean (SD)** | **P value** |
| --- | --- | --- | --- |
| Gender |  |  |  |
| Male | 678 (47.7) | 3.5 (1.1) | 0.795 |
| Female | 740 (52.1) | 3.5 (1.1) |  |
| Other* | 2 (0.2) | 2.5 (0.7) |  |
| Age (years) |  |  |  |
| 18-29 | 295 (20.8) | 3.5 (1.1) | Ref |
| 30-49 | 508 (35.8) | 3.6 (1.1) | 0.147 |
| 50-69 | 419 (29.5) | 3.4 (1.1) | 0.865 |
| 70+ | 198 (13.9) | 3.4 (1.1) | 0.880 |
| Aboriginal and/or Torres Strait Islander |  |  |  |
| Yes | 47 (3.3) | 3.8 (1.2) | **0.032** |
| No | 1373 (96.7) | 3.5 (1.1) |  |
| Country of birth |  |  |  |
| Australia | 1096 (77.2) | 3.5 (1.1) | 0.068 |
| Other | 324 (22.8) | 3.6 (1.1) |  |
| Employment status |  |  |  |
| Not working | 591 (41.6) | 3.3 (1.2) | **<0.001** |
| Working full/part time | 829 (58.4) | 3.6 (1.1) |  |
| Educational attainment |  |  |  |
| Year 10 or below | 161(11.3) | 3.3 (1.3) | Ref |
| High school | 235 (16.5) | 3.3 (1.1) | 0.253 |
| Trade/apprenticeship/cert | 483 (34.0) | 3.4(1.1) | **0.019** |
| University degree | 541 (38.1) | 3.6 (1.1) | **0.009** |
| Children in household |  |  |  |
| Attending childcare/school | 212 (14.9) | 3.4 (1.1) | **<0.001** |
| Not attending childcare/school or no children | 1208 (85.1) | 3.8 (1.1) |  |
| Travelled internationally in 2020 |  |  |  |
| Yes | 222 (15.6) | 3.7 (1.1) | **0.001** |
| No | 1198 (84.4) | 3.4 (1.1) |  |
| Have private health insurance |  |  |  |
| Yes | 830 (58.5) | 3.5 (1.1) | **0.01** |
| No | 590 (41.5) | 3.4 (1.2) |  |
| Health rating |  |  |  |
| Very good/good | 1009 (71.1) | 3.5 (1.1) | 0.823 |
| Moderate | 294 (20.7) | 3.5 (1.1) | 0.420 |
| Poor/very poor | 117 (8.2) | 3.6 (1.3) | Ref |
| Chronic health condition |  |  |  |
| Present | 363 (25.6) | 3.7 (1.1) | **<0.001** |
| None | 1057 (74.4) | 3.4 (1.1) |  |

\*Not included in comparison due to small numbers.
Numbers in bold are statistically significant.

Eighty percent (n=1143) agreed with the statement that *getting myself vaccinated for COVID-19 would be a good way to protect myself against infection*, while a further 194 (13%) were uncertain, leaving 83 (5.8%) to disagree with the sentiment (Table 2). Beyond individual perceptions, participants were asked to comment on perceived support from family and friends towards receipt of a COVID-19 vaccine, of which 1118 (78%) agreed that they would be supported. A similar level of support was given to the statement ‘to protect the health of the community, we should follow government guidelines about vaccines’ with 1190 (84%) agreeing.

**Table 2.** **Perceptions towards vaccination in general and the COVID-19 vaccine**.

| **Question** | **Agree (%)** | **Disagree (%)** | **Neutral** |
| --- | --- | --- | --- |
| Vaccines are effective at preventing diseases. | 1188 (83.7) | 69 (4.9) | 163 (11.5) |
| Diseases provide better immunity than vaccines do. | 305 (21.5) | 550 (38.7) | 565 (39.8) |
| I generally do what my health care professional recommends | 1195 (84.2) | 62 (4.4) | 163 (11.5) |
| Getting myself vaccinated for COVID-19 would be a good way to protect myself against infection | 1143 (80.5) | 83 (5.8) | 194 (13.7) |
| My family and friends would probably think that getting a COVID-19 vaccine is a good idea. | 1118 (78.7) | 93 (6.5) | 209 (14.7) |
| To protect public health, we should follow government guidelines about vaccines. | 1190 (83.8) | 62 (4.1) | 168 (11.8) |
| Patients with risk factors should be the first ones to get the COVID-19 vaccine when available. | 1211 (85.3) | 58 (4.1) | 151 (10.6) |
| Healthcare workers should be the first ones to get the COVID-19 vaccine | 1198 (84.4) | 51 (3.6) | 171 (12.0) |

When it came to prioritization of target groups for a future COVID-19 vaccine, participants were strongly in favour of healthcare workers being the first ones to get the vaccine (1198, 84%). Only 51 (3.5%) participants disagreed with that sentiment, while the remaining participants were neutral (171/1420, 10%). The same level of support was shown to the prioritization of patients with risk factors, with 1211 (85%) agreeing that they should be the first ones to get the COVID-19 vaccine. Again only 58 (4%) participants disagreed. Interestingly, there was equal distribution across age groups and chronic health conditions for both variables.

The median score for risk perception of COVID-19 infection amongst those who would not accept the vaccine was 3 (IQR: 2– 4) compared to a median of 4 (IQR: 3-4) among those who would accept the vaccine (p < 0.001). Mean risk perception scores was significantly higher among Aboriginal and/or Torres Strait Islander participants (P=0.032) compared to non-Indigenous participants, those who were working full time/part time (P<0.001) compared to unemployed people. Participants who had a trade/apprenticeship/certificate or a University degree had significantly higher mean risk score compared to participants with educational level year 12 or below (P=0.019 and P=0.009 respectively). Similarly, respondents having private health insurance (P=0.01) and those with chronic health conditions (P=?) perceived their mean risk score higher than those without.

There was variation in the proportion of people who agreed that getting *vaccinated against COVID-19 would be a good way to protect myself against infection* by demographic characteristics. These differences were significant for gender, Indigenous status, educational attainment, private health insurance, international travel in 2020 and self-reported chronic health condition (Table 3). Overall, 83% of females agreed with the statement compared to 78% of males (aOR=1.4 (95% CI: 1.1-1.8); P=0.029. Those above 70 years of age (90.9%) compared to those between 18-29 years of age (76.6%) reported higher level of agreement (aOR=2.3 (95% CI 1.2-4.1); P=0.008) Agreement was also higher for those who self-reported having a chronic disease (aOR=1.4 (95% CI: 1.1-2.0); P=0.043) and who had private health insurance (aOR=1.7 (95% CI: 1.3-2.3); P<0.001) (Table 3).

**Table 3:** **Univariate analysis and multivariate logistic regression model of Covid-19 vaccine acceptance and demographic variables**.

|  | **Covid-19 vaccine acceptance n (%)** | **Unadjusted ORs (95% CI)** | **P value** | **Adjusted ORs (95% CI)** | **P value** |
| --- | --- | --- | --- | --- | --- |
| Gender |  |  |  |  |  |
| Male | 529 (78.0) | REF |  | REF |  |
| Female | 614 (83.0) | **1.4 (1.1-1.8)** | **0.019** | **1.4(1.1-1.8)** | **0.029** |
| Other* | 0 (0.0) | -- |  | -- |  |
| Age (years) |  |  |  |  |  |
| 18-29 | 226 (76.6) | REF |  | REF |  |
| 30-49 | 401 (78.9) | 1.1 (0.8-1.6) | 0.442 | 1.0 (0.7-1.3) | 0.899 |
| 50-69 | 336 (80.2) | 1.2 (0.9-1.8) | 0.250 | 1.0 (0.7-1.5) | 0.844 |
| 70+ | 180 (90.9) | **3.1 (1.8-5.3)** | **<0.001** | **2.3 (1.2-4.1)** | **0.008** |
| Aboriginal and/or Torres Strait Islander |  |  |  |  |  |
| Yes | 32 (68.1) | **2.0 (1.1 -3.7)** | **0.032** | 0.7 (0.3-1.3) | 0.202 |
| No | 1111 (80.9) | REF |  | REF |  |
| Country of birth |  |  |  |  |  |
| Australia | 871 (79.5) | REF | 0.075 | 1.3 (0.9-1.8) | 0.179 |
| Other | 272 (84.0) | 1.4 (1.0-1.9) |  |  |  |
| Employment status |  |  |  |  |  |
| Not working | 488 (82.6) | REF | 0.095 | REF | 0.775 |
| Working full/part time | 655 (79.0) | 0.8 (0.6-1.0) |  | 0.9 (0.7-1.3) |  |
| Educational attainment |  |  |  |  |  |
| Year 10 or below | 122 (75.8) | REF |  | REF |  |
| High school | 192 (81.7) | 1.4 (0.9-2.3) | 0.154 | 1.5 (0.9-2.5) | 0.124 |
| Trade/apprenticeship/cert | 396 (82.0) | 1.5 (0.9-2.2) | 0.086 | 1.5 (1.0-2.4) | 0.076 |
| University degree | 433 (80.0) | 1.3 (0.8-1.9) | 0.244 | 1.3 (0.8-2.1) | 0.254 |
| Employment status |  |  |  |  |  |
| Not working | 488 (82.6) | REF |  | REF |  |
| Working full/part time | 655 (79.0) | 0.8(0.6-1.0) | 0.095 | 1.0 (0.7-1.3) | 0.849 |
| Children in household |  |  |  |  |  |
| Attending childcare/school | 167 (78.8) | 0.9 (0.6-1.3) | 0.494 | Not included in the model |  |
| Not attending childcare/school or no children | 976 (80.8) | REF |  |  |  |
| Travelled internationally in 2020 |  |  |  |  |  |
| Yes | 166 (74.2) | REF |  | REF |  |
| No | 977 (81.6) | **1.5 (1.1-2.1)** | **0.020** | 1.5 (1.0-2.1) | 0.05 |
| Have private health insurance |  |  |  |  |  |
| Yes | 693 (83.5) | **1.6 (1.2-2.0)** | **0.001** | **1.7 (1.3-2.3)** | **<0.001** |
| No | 450 (76.3) | REF |  | REF |  |
| Health rating |  |  |  |  |  |
| Very good/good | 809 (80.2) | 1.1 (0.7-1.8) | 0.692 | Not included in the model |  |
| Moderate | 242 (82.3) | 1.3 (0.7-2.2) | 0.389 |  |  |
| Poor/very poor | 92 (78.6) | REF |  |  |  |
| Chronic health condition |  |  |  |  |  |
| Present | 307 (84.6) | **1.4 (1.1-2.0)** | **0.024** | **1.4 (1.1-2.0)** | **0.043** |
| None | 836 (79.1) | REF |  | REF |  |

Values in bold are statistically significant (P<0.05)

## DISCUSSION

The survey was conducted in March 2020, at a time when the first wave of COVID-19 cases was increasing in Australia, there was intense media coverage and community members were being encouraged to adopt hygiene and physical distancing strategies. At that point, there was no lockdown enforced in Australia. From our survey, we found that 80% agreed that receiving the COVID-19 vaccine would be a good way to protect themselves. The level of agreement amongst our participants varied in comparison to other studies. An online survey of the French population (conducted 10 days after the nationwide lockdown was introduced in March) found that 74% would use a vaccine [8]. A similar acceptance rate was reported in other surveys (conducted around March-April) of residents in the United States (67%-69%) [9, 10], Indonesia (67% to 95% depending on the effectiveness of the vaccine) [11] and 73% for parts of Europe (Denmark, France, Germany, Italy, Portugal, the Netherlands, and the UK) [12]. The difference in acceptance rate documented in this study may be due to a single or combination of factor(s) including: (1) the variation in the wording of the question; (2) high level of confidence and trust in the Australian government [6] or (3) due to concerns about increasing local transmission which were high at the time. However, our results align with other Australian studies, which have reported willingness levels between 76% to 86% [13, 14]. Both studies collected the data in April 2020.

It has been well documented that the same psychological factors that influence acceptance of national immunisation program vaccines apply during pandemics [15]. Studies conducted in 2009 examining the acceptance of the pandemic influenza A/H1N1 vaccine found that perceptions of risk and severity played a key role in whether people agreed with the necessity of vaccination [4, 16]. At the time that the H1N1 pandemic immunisation programs were commenced in Australia, it was well after the peak of the pandemic (which was already deemed as ‘moderate’ by governments and other agencies). This impacted the perceived personal risk of infection, as well as how people perceived the severity of the infection, which resulted in low levels of vaccine uptake [4, 17-19]. While the characteristics of the COVID-19 pandemic are vastly differently to the H1N1 influenza pandemic in 2009, it is important that we consider how we are going to engage and communicate with those in the community who perceive their personal risk as low. In mid-March, we identified that 74% of our study participants ranked their personal risk of acquiring COVID-19 as ‘intermediate’ to ‘very-low’ [6]. With this group, it may be necessary to draw on the influence of anticipated regret, which has been found to be an important determinant of intention to vaccinate [20, 21]. While the expectation of anticipated regret is primarily cognitive, it also likely has an affective component, as imagining an unpleasant future may elicit emotion in the present [22]. People may act to reduce what they expect to experience by acting. Examples could be: (1) anticipated regret of not getting the COVID-19 vaccine, as a family member gets infected, encourages vaccination; and (2) anticipated regret of not getting the COVID-19 vaccine, as a person is unable to travel abroad to visit friends and relatives (hypothetical situation of COVID-19 vaccination operating in the same manner as yellow fever vaccination), which encourage vaccination.

To translate early willingness into actual vaccine receipt, we will need to draw on key behavioural insights from past studies. For example, a recommendation from a healthcare provider is a key driver of routine immunisation uptake [23-26]. Amongst our participants, the majority agreed that they follow the advice of their healthcare professionals. To support this action, there is a need to equip healthcare professionals with the understanding about the COVID-19 vaccine (including how it was developed, safety profiles), the skills to take a presumptive approach to recommending the vaccine and the confidence to answer questions. For example, there may be a need to support peoples understanding around the rational for receiving the COVID-19 vaccine, especially amongst those who believe that they may have been already infected during the pandemic. Around a quarter of our participants agreed with the statement that ‘diseases provide better immunity than vaccines do’, while a further 40% were neutral about the statement. Health professionals will have a strong impact on uptake since they both recommend, and in this case, are likely to be the first eligible for the vaccine.

In settings like Australia where vaccines are delivered predominately in primary care settings, the focus will be on supporting General Practitioners and Practice Nurses. However, given the adult risk groups likely targeted with a COVID-19 vaccine, other providers will need to be considered. For example, hospital and private practice specialists (medical and nursing) may be a trusted source of information about the COVID-19 vaccine for those people with chronic medical conditions [27]. There may be high levels of confidence in vaccine information being provided by these specialists, as they are experts in a specific chronic medical condition [27]. This may be especially important if the vaccine has any contraindications or precautions for people with any chronic conditions or who are immunosuppressed. There may be other providers that need to be supported to effectively communicate about this vaccine. Given that not all adults regularly connect with primary care, there will be a need to support community-controlled health organisations to promote uptake among their local communities. Public health campaigns may also need to consider enlisting other partners, outside of traditional medical and public health communities, to support activities that promote awareness and acceptance of the vaccine. These may include peak bodies which are not-for-profit non-government health-condition specific organisations that focus on one health condition/disease and disseminate evidence-based information related to their conditions and health [28]. Information delivered by these groups would be relevant and credible to their constituents.

Populations at risk of COVID-19 infection are diverse in social, behavioural, cultural and health practices as well as their understanding of COVID-19. Racial and ethnic disparities in the severity of COVID-19 illness have been identified [29]. In non-pandemic periods, people from culturally and linguistically diverse (CALD) backgrounds can be disadvantaged by the factors that contribute to health inequity and have been documented as resulting in lower uptake of recommended vaccines including influenza [30]. To support access to this vaccine and equity in the delivery, it is critical that engagement approaches are tailored so they meet the needs of all communities, in terms of messages and vaccine dissemination strategies [31]. For example, communicating messages about the vaccine to CALD communities is not just a question of providing translations of information that meet readability assessment scores.

Previously, Mileti and Darlington (1997) found that people from CALD backgrounds generally prioritize social networks and interpersonal communication when seeking information and prefer to receive information from people with similar attributes as themselves [32]. There will be a need to involve community leaders with the promotion of a vaccine including cultural and religious leaders, and Aboriginal elders. Use of these community influencers may support the engagement of Aboriginal communities and CALD groups including newly arrived migrants who rely on informal information sources through social networks and particularly in early stages of settlement [33]. These actors may have heightened success in delivering relevant culturally appropriate messages via formats and venues which may not be reached by mainstream mass communication approaches. Beyond ensuring that messages are effectively disseminated into all parts of the community, there is also a need to think about access in terms of convenience, location of vaccine services, and time-costs associated with receive it. There may be a need to think beyond primary care to reduce access barriers for some communities [34].

When the first trials commenced in the US, rumours began to circulate that fake vaccines were being used, while in the UK, the first subjects enrolled in vaccine trials were forced to clarify that they were still alive [35]. Mis- and disinformation is going to continue to circulate during this pandemic and will surge with the availability of COVID-19 vaccines. To respond to the “infodemic”, the WHO put together a framework based on a crowdsourcing exercise to support governments to manage the issue [36]. The work culminated in six key principles that governments could start to consider when planning their activities around the promotion and delivery of the COVID-19 vaccine. One key area highlighted was the need to slow down and streamline the flow of information of all kinds. Having transparent information, which is adapted to local languages, literacy levels, is regularly updated, and focuses on common/known mild reactions to new vaccines may assist with stemming the flow of misinformation about the safety of the vaccines [36]. The importance of this was identified in 2009 by Eastwood et.al who reported a critical link between willingness to accept a pandemic H1N1 vaccine and the availability of easily interpretable vaccine safety data [5].

The strengths of our study include a large, representative cross-section of the adult Australian population. However, the work is subject to several limitations including that we recruited a convenience sample of participants. People who could not communicate in English were excluded from the sample, which may have affected representation of ethnic minorities. We also had under-representation of Aboriginal and Torres Strait Islander peoples and those residing in remote settings. As participation in our study was on a voluntary basis, this study has potential for self-selection bias by community members who are particularly concerned about this pandemic.

## CONCLUSION

Throughout this pandemic, there have been issues with communication, shifts in recommendations and fluctuations in cases, which all have the potential to undermine trust in governments. To support an effective launch of new COVID-19 vaccines, governments need to understand the community’s concerns, and identify strategies that will support engagement. There is a pressing and critical need to start planning public health communication strategies that are designed to support healthcare professionals and those in civil society who may play a role, as well as engage all members of the community.

## Data Availability

The dataset used and/or analysed during the current study are available from the corresponding author on reasonable request

## List of abbreviations

None

## DECLARATIONS

### Ethics approval and consent to participate

Ethics approval for the study was obtained from the University of New South Wales HREAP G: Health, Medical, Community and Social (HC200190). After reading the participant information, consent was implied if the person completed the survey and submitted it via the QOR website

### Consent for publication

Not applicable

### Competing interests

Dr Holly Seale has previously received funding from drug companies for investigator driven research and consulting fees to present at conferences/workshops and develop resources (bio-CSL/Sequiris, GSK and Sanofi Pasteur). She has also participated in advisory board meeting for Sanofi Pasteur. She is a Section Editor for BMC Infectious Diseases. The other authors do not have anything to declare.

### Funding

There was no funding associated with this study

### Authors’ Contributions

HS conceived the study, undertook the data collection, and developed the journal paper. All other authors assisted with the design, interpretation and with revising the paper. All authors have read and approved the manuscript.

## Acknowledgements

We would like to thank the respondents for their time in participating in the research study.

## Notes

### Author Declarations

Ethics approval for the study was obtained from the University of New South Wales HREAP G: Health, Medical, Community and Social (HC200190).

